# Comparison of psychological distress and demand induced by COVID-19 during the lockdown period in patients undergoing peritoneal dialysis and hemodialysis: a cross-section study in a tertiary hospital

**DOI:** 10.1101/2020.04.13.20063099

**Authors:** Xiaoxiao Xia, Xiaofang Wu, Xueli Zhou, Zhiyun Zang, Li Pu, Zi Li

## Abstract

**Background:** Since the outbreak of COVID-19 in December 2019, it has spread rapidly and widely, bringing great psychological pressure to the public. In order to prevent the epidemic, lockdown was required in many areas of China, which led to inconvenience of treatment for dialysis patients. To explore the psychological distress and the psychological demand induced by COVID-19 in the patients undergoing dialysis and compare the difference between hemodialysis (HD) and peritoneal (PD) patients during the lockdown period.

**Methods:** Questionnaires were given to the dialysis patients in West China Hospital of Sichuan University. The Impact of Event Scale (IES) was used to investigate the patients’ trauma-related distress in response to COVID-19.

**Results:** 232 eligible respondents were enrolled in this cross-section study, consisting of 156 PD patients and 76 HD patients. The median IES score for all the enrolled patients was 8.00 (2.00-19.00), which belonged to the subclinical dimension of post-traumatic stress symptoms. HD patients had a significant higher IES score than PD patients (11.50 vs 8.00) (p<0.05). HD patients already got more psychological support from the medical staff. There was no significant difference on further demand of psychological support between the two groups. In the multivariate regression analysis, we found that dialysis vintage, the impact of COVID-19 on the severity of illness and daily life, and confidence in overcoming the disease contributed to IES score (p<0.05).

**Conclusions:** HD patients had more severe trauma-related stress symptoms than PD patients. When major public healthy events occurred, careful psychological estimate and sufficient psychological support should be provided to the dialysis patients, especially to the HD patients.

## INTRODUCTION

In December 2019, the Coronavirus Disease 2019 (COVID-19) broke out in Wuhan, China [1]. Then this disease spread quickly to other provinces in China and some other countries [2]. Till February 29, 2020, China has reported a total of 79,394 confirmed cases of COVID-19, including 2,838 deaths [3]. As for the transmission of COVID-19, human-to-human transmission has been confirmed [4]. The spreading of the virus can occur through coughing, sneezing and talking because of the droplets they produce [5]. The epidemic of COVID-19 had brought great pressure to the public [6].

It’s reported that after public health emergencies, the prevalence of depression varies from 5.4% to 52%, and the suicide rate also shows an upward trend [7], which means that such health emergencies have great psychological impact to the population. After the health emergencies, such as Severe Acute Respiratory Syndrome (SARS) epidemic in 2003, the Ebola outbreak in 2014, the Middle East Respiratory Syndrome (MERS) period in 2015 and so on, psychological problems were reported, such as fear, boredom, anxiety and depression [8-11]. And several studies reported that taking effective psychological intervention was essential to improve the mental health of the population after the epidemic [12,13]. Research observed that for dialysis patients, 38.1% of them had symptoms including anxiety and depression and 57.1% presented stress [14] even under normal circumstances. But little is known about the mental health of hemodialysis patients (HD) or peritoneal dialysis (PD) patients in the context of public health emergencies.

Due to the human-to-human transmission of COVID-19, strict prevention and control solutions were required, which brought many problems to dialysis patients. Firstly, in order to prevent the spreading, the Chinese government initiated first-level responses to major public health emergencies and the public transport was suspended [15]. People were limited within their community, which made it difficult for some of the dialysis patients to go to the hospital. Secondly, there was a sharp increase of febrile and suspected cases in the hospitals designated to receive such patients [16]. Therefore, the hospitals were high-risk areas for cross infection, but the dialysis patients had to go there for treatment, especially the hemodialysis patients. Unfortunately, the patients with end-stage renal disease (ESRD) are susceptible to infection because of low immunity [17]. Thirdly, the shortage of protective materials caused the psychological panic or anxiety of ordinary people, including people outside Hubei Province. Altogether, the outbreak of COVID-19 may bring great psychological pressure to the dialysis patients.

Dialysis is an alternative treatment for ESRD patients in addition to renal transplantation. Peritoneal dialysis is a home-based dialysis method, which has the advantages of convenience and does not need special medical instruments [18]. Therefore, many PD patients can undergo peritoneal dialysis at home by themselves and don’t need to go to the hospital frequently. But, the HD patients have to go to dialysis center for treatment 2-3 times a week [19]. It’s reported that with the rapid increase of confirmed cases and deaths of COVID-19, the public has been experiencing psychological problems [15]. However, there are few studies on the mental health of dialysis patients with ESRD after health emergencies and the comparison between HD and PD is less. We intend to conduct this study to explore the psychological distress and the psychological demands induced by COVID-19 in the patients undergoing dialysis and compare the difference between HD and PD patients during the lockdown period.

## MATERIALS AND METHODS

### Study Design and Population

Questionnaires were given to our dialysis patients from February 24 to February 29, 2020 and conducted by smart phones. The patients included all the PD patients who followed up regularly in the department of nephrology and the HD patients in Wenjiang branch, both of them belong to the West China Hospital of Sichuan University. The inclusion criteria were dialysis patients with ESRD over 18 years old and could use the smart phones to fulfill the questionnaires, consent was obtained before the data collection. Those who could not use smart phones or were unwilling to answer the questionnaires were excluded.

The questionnaire consisted of four parts as basic demographic data, the impact of COVID-19 on the severity of illness and daily life, the Impact of Event Scale (IES) and their psychological demands during the epidemic.

### Impact of Event Scale

The Impact of Event Scale is a self-report scale that has been used widely to investigate trauma-related distress in response to a specific stressful life event and has demonstrated extensive reliability and validity [20,21]. Each of the 15 items is rated on a four-point frequency scale (0, not at all; 1, rarely; 3, sometimes; 5, often). The IES yields a total score (ranging from 0 to 75) and subscales scores, which can be calculated for the intrusion (ranging from 0 to 35) and avoidance (ranging from 0 to 40) [22]. The total IES scores can be interpreted according to the following dimensions of post-traumatic stress symptoms: 0 to 8 (subclinical range), 9 to 25 (mild range), 26 to 43 (moderate range), 44+ (severe range). It is suggested that the cut-off point is 26, above which a moderate or severe impact is indicated, and psychological referral is suggested [22].

### Statistical Analyses

Statistical analysis was done with SPSS (version 21.0). Continuous variables were expressed as means ± SDs, or medians (interquartile ranges). Categorical variables were expressed as number and percentages (%). Student t test or Mann-Whitney U test were used for continuous variables and chi-square test was used for categorical variables. The unitary linear correlation was used to examine the relationship between IES scores and other variables, and then the significant factors were further analyzed for IES score using multivariate regression analysis. A two-tailed p < 0.05 was considered statistically significant.

## RESULTS

### Patient characteristics

Of the 254 questionnaires received in the survey, 22 patients were excluded, among which fifteen patients (5.91%) had incomplete questionnaires and seven patients (2.76%) underwent combined PD and HD. The remaining 232 respondents included 156 PD patients and 76 HD patients in Wenjiang branch. The successful respondents comprised 104 (44.83%) male and 128 (55.17%) female, with a mean age of 44.21 years (range 22-85 years). PD group had a higher percentage of newly initiation of dialysis (less than 1 year) compared to HD group (27.6% vs 7.9%), while HD group had a higher proportion of dialysis vintage more than 5 years than PD group (32.9% vs 19.9%). HD patients had an obviously longer dialysis vintage than the PD patients (P<0.01). Most of the HD patients (86.8%) lived in Chengdu, where our hospital located. About 50.6% PD patients lived in other areas outside Chengdu (P<0.01). There were no significant differences in gender, age, education, marital status, or occupation between the two groups (Table 1).

**Table 1.** Patient Characteristics.

|  | **HD**<br>**(N=76)** | **PD**<br>**(N=156)** | **Total**<br>**(N=232)** | **P value** |
| --- | --- | --- | --- | --- |
| **Gender, n (%)** |  |  |  | 0.067 |
| Male | 41 (53.9) | 63 (40.4) | 104 |  |
| Female | 35 (46.1) | 93 (59.6) | 128 |  |
| **Age (years), n (%)** |  |  |  | 0.378 |
| ≤40 | 33 (43.4 ) | 65 (41.7) | 98 |  |
| 41-60 | 41 (53.9 ) | 74 (47.4) | 115 |  |
| ≥61 | 2 (2.6) | 17 (10.9) | 19 |  |
| **Marital status, n (%)** |  |  |  | 0.473 |
| Married | 59 (77.6) | 123 (78.8) | 182 |  |
| Single | 7 (9.2 ) | 20 (12.8) | 27 |  |
| Divorced | 9 (11.8) | 10 (6.4 ) | 19 |  |
| Widowed | 1 (1.3) | 3 (1.9 ) | 4 |  |
| **Education, n (%)** |  |  |  | 0.162 |
| Primary | 4 (5.3) | 8 (5.1) | 12 |  |
| Junior | 20 (26.3) | 60 (38.5) | 80 |  |
| Senior | 25 (32.9) | 41 (26.3) | 66 |  |
| University degree or above | 27 (35.5) | 47 (30.1) | 74 |  |
| **Occupation, n (%)** |  |  |  | 0.473 |
| Medical staff | 2 (2.6) | 3 (1.9) | 5 |  |
| Worker/farmer | 15 (19.7) | 42 (26.9) | 57 |  |
| Teacher | 5 (6.6 ) | 6 (3.8) | 11 |  |
| Government | 5 (6.6 ) | 5 (3.2) | 10 |  |
| Company employee | 7 (9.2 ) | 11 (7.1) | 18 |  |
| Retired | 8 (10.5) | 26 (16.7) | 34 |  |
| Unemployment or others | 34 (44.7 ) | 63 (40.4) | 97 |  |
| **Habitation, n (%)** |  |  |  | 0.000 |
| City of Chengdu | 66 (86.8) | 77 (49.4) | 143 |  |
| Other areas outside Chengdu | 10 (13.2) | 79 (50.6) | 89 |  |
| **Dialysis vintage (years), n (%)** |  |  |  | 0.000 |
| <1 | 6 (7.9) | 43 (27.6) | 49 |  |
| 1-2 | 27 (35.5) | 55 (35.3) | 82 |  |
| 3-5 | 18 (23.7) | 27 (17.3) | 45 |  |
| >5 | 25 (32.9) | 31 (19.9) | 56 |  |

### Comparisons of the impact on the severity of illness and daily life

In our study, most of HD patients (94.7%) needed to visit the hospital three or more times per week. On the contrary, PD patients could do dialysis at home by themselves, and 80.1% of patients go to the hospital since the outbreak (P<0.01). The family members of some PD patients (30.1%) could go to the hospital for prescription of medications and PD fluid instead. Most of the patients didn’t feel that COVID-19 had obvious impact on the severity of illness or daily life and there was no significant difference between the two groups (Table 2). Only 17.1% HD patients thought that COVID-19 had moderate or more severe influence on the severity of their illness, and only 13.5% PD patients did so. As for the impact on the daily life, 26.3% HD patients and 21.2% PD patients thought that COVID-19 had a moderate or more impact on their daily life, respectively. As to the reasons of going out, dialysis treatment was the main reason for HD patients, while prescribing medicine and shopping for necessities were the main reasons for PD patients (Table 2). What’s when being asked about the most wanted supports, HD patients mainly wanted the hospitals remained open and protective equipment were adequate, which were essential for hemodialysis treatment. On the other side, PD patients hoped that the delivery of medication and PD fluid could be more convenient and the hospitals remained open as well (Table 2).

**Table 2.** Comparisons between HD and PD on the impact of COVID-19 to the severity of illness and daily life.

|  | **HD**<br>(N =76) | **PD**<br>(N =156) | **Total**<br>(N =232) | **P value** |
| --- | --- | --- | --- | --- |
| **Frequency to hospital per week, n (%)** |  |  |  | 0.000 |
| 0 | 0 (0.0) | 125 (80.1) | 125 |  |
| 1-2 | 4 (5.3) | 28 (17.9) | 32 |  |
| ≥3 | 72 (94.7) | 3 (1.9 ) | 75 |  |
| **Influence on severity of illness, n (%)** |  |  |  | 0.370 |
| No | 34 (44.7) | 53 (34.0) | 87 |  |
| Mild | 29 (38.2) | 82 (52.6) | 111 |  |
| Moderate | 9 (11.8) | 17 (10.9) | 26 |  |
| Severe | 4 (5.3) | 4 (2.6) | 8 |  |
| **Influence on daily life, n (%)** |  |  |  | 0.402 |
| No | 14 (18.4) | 31 (19.9) | 45 |  |
| Mild | 42 (55.3) | 92 (59) | 134 |  |
| Moderate | 13 (17.1) | 26 (16.7) | 39 |  |
| Severe | 7 (9.2) | 7 (4.5) | 14 |  |
| **Who went to the hospital, n (%)** |  |  |  | 0.000 |
| Only myself | 69 (90.8) | 87 (55.8) | 156 |  |
| Replaced by family Members | 0 (0.0) | 47 (30.1) | 47 |  |
| Accompanied by family members | 7 (9.2) | 22 (14.1) | 29 |  |
|  | **HD, n (%)** | **PD, n (%)** | **Responses**<br>**n (%)** | **Percent of**<br>**Cases (%)** |
| **Reasons of going out** |  |  |  |  |
| Total | 76 (100) | 156 (100) | 321 (100.0) | 138.4 |
| Shopping | 32 (42.1) | 80 (51.3) | 112 (34.9) | 48.3 |
| Therapy | 71 (93.4) | 26 (16.7 ) | 97 (30.2) | 41.8 |
| Prescribe medicine | 6 (7.9) | 66 (42.3 ) | 72 (22.4 ) | 31.0 |
| Work | 1 (1.3) | 13 (8.3 ) | 14 (4.4) | 6.0 |
| Walk | 0 (0.0) | 17 (10.9) | 17 (5.3) | 7.3 |
| Others | 0 (0.0) | 9 (10.9) | 9 (2.8) | 3.9 |
| **Supports wanted** |  |  |  |  |
| Total | 6 (100) | 156 (100) | 524 (100) | 225.9 |
| Protective equipment from government | 45 (59.2) | 62 (39.7) | 107 (20.4) | 46.1 |
| Hospitals remained open | 63 (82.9) | 71 (45.5) | 134 (25.6) | 57.8 |
| Convenient drug delivery | 8 (10.5) | 93 (59.6) | 101 (19.3) | 43.5 |
| Protective information from medical staff | 14 (18.4) | 11 (7.1) | 25 (4.8) | 10.8 |
| Adjust the treatment protocol | 4 (5.3) | 23 (14.7) | 27 (5.2) | 11.6 |
| Strengthen government management | 39 (51.3) | 53 (34.0) | 92 (17.6) | 39.7 |
| Family support | 8 (10.5) | 11 (7.1) | 19 (3.6) | 8.2 |
| Others | 2 (2.6) | 15 (9.6) | 17 (3.2) | 7.3 |

### Comparisons of psychological supports and demands

The two groups had significant difference on the psychological support received from medical staff (p<0.05). More than half of HD patients (55.3%) admitted receiving great psychological support from medical staff, and only 1 patient (1.3%) complained that he didn’t receive any support. For PD patients, 39.7% of them admitted receiving great support, and 16.0% received no support from medical staff at all. More HD patients were informed on how to protect themselves by the medical staff than PD patients (100% vs 74.4%) (P<0.05) (Table 3).

**Table 3.**
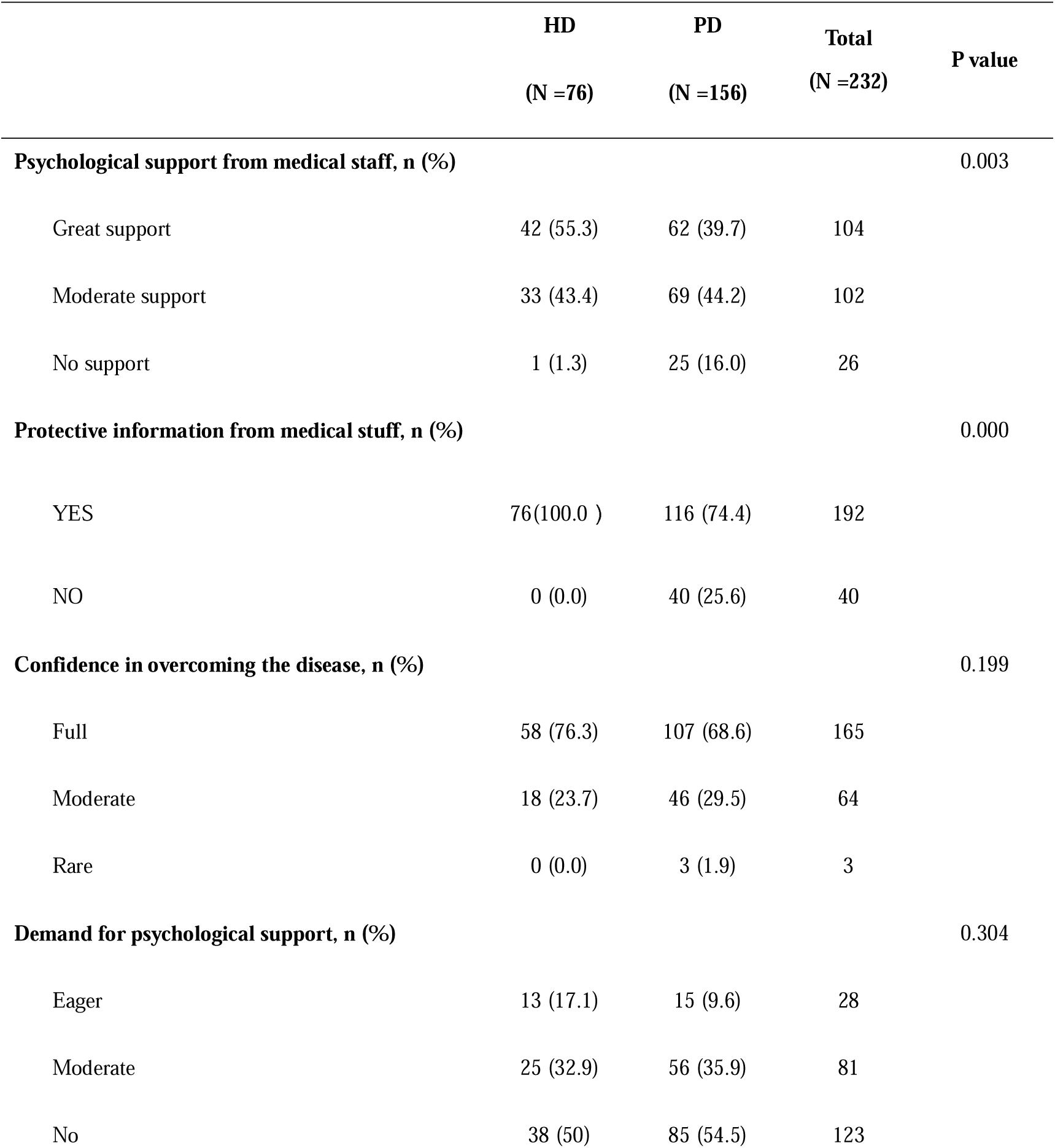
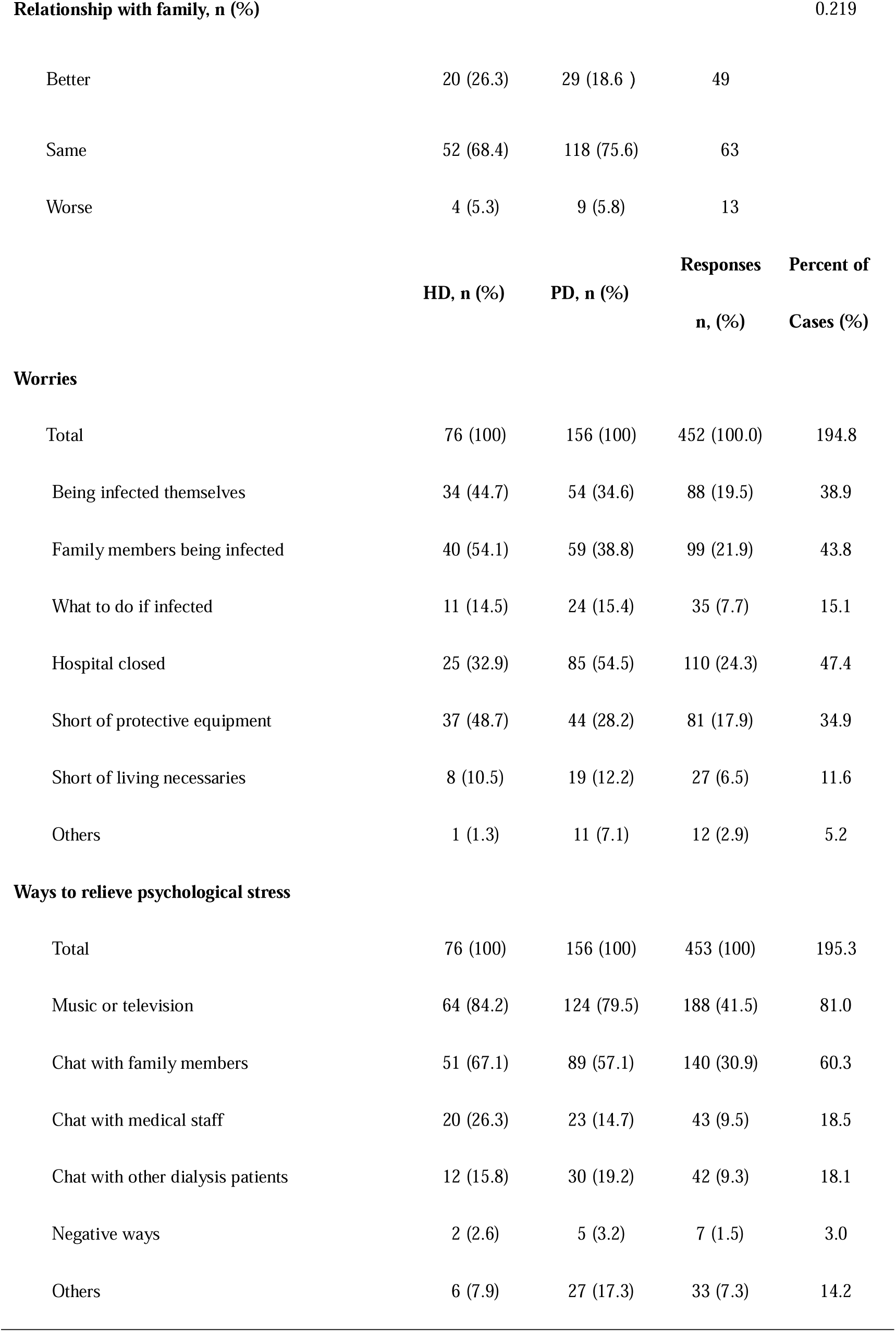
Comparisons between HD and PD on psychological support.

During the period being limited within their communities, both groups relieved their psychological distress mainly by music, television or chatting with family members, and rarely chose negatives ways. What’s more, HD patients seemed more likely to choose chatting with medical staff for psychological support compared with PD patients (26.3% vs 14.7%). No significant difference was found between the two groups on the relationship with their family. There was also no significant difference on the further demand of psychological support between the two groups. Only a small number of patients in both groups expressed a strong desire for psychological support (17.1% in HD patients and 9.6% in PD patients) (P>0.05). And the majority of the patients have full confidence in overcoming the disease (76.3% of HD patients vs 68.6% of PD patients) (P>0.05) (Table 3).

Both of the two groups had some patients worried about themselves (44.7% in HD vs 34.6% in PD) or their family members (54.1% in HD vs 38.8% in PD) being infected by COVID-19. We also found that HD patients had more worries about the shortage of protective equipment, while PD patients worried more about the hospital might be closed (Table 3).

### IES score and the severity of psychological distress

The median IES score for all the enrolled patients was 8.00 (2.00-19.00), which belonged to the subclinical dimension of post-traumatic stress symptoms. HD patients had a significant higher score than PD patients (11.50 vs 8.00) (p<0.05). The discrepancy mainly lied in avoidance symptoms, with the median score as 6.00 in HD patients and 3.50 in PD (p<0.05) (Table 4). What’s more, the severity of stress symptoms varied between the two groups (p<0.05). In the HD group, 43.4% patients were subclinical, 34.2% were mild and 22.4% were moderate or severe. However, in the PD group, more patients were subclinical (57.1%), 29.5% were mild and 13.4% were moderate or severe (Table 4).

**Table 4.** Comparisons between HD and PD on IES.

|  | HD | PD | P value |
| --- | --- | --- | --- |
| **IES scores: median (IQR\*)** |  |  |  |
| IES (Total) | 11.50 (3.00-25.00) | 8.00 (1.00-15.00) | 0.02 |
| Avoidance | 6.00 (1.00-13.75) | 3.50 (0-8.75) | 0.023 |
| Intrusion | 4.50 (1.25-12.00) | 3.00 (0.25-7.00) | 0.062 |
| **Severity, n (%)** |  |  |  |
| Subclinical (0-8 points) | 33 (43.4) | 89 (57.1) | 0.036 |
| Mild (9-25 points) | 26 (34.2) | 46 (29.5) |  |
| Moderate (26-43 points) | 12 (15.8) | 13 (8.3) |  |
| Severe ($\geq 44$ points) | 5 (6.6) | 8 (5.1) | |

\*IQR: Inter-quartile range

### Univariate analysis and multivariate analysis: risk of IES score

After conducting the univariate analysis, our study revealed that for all the enrolled patients, the median IES score was significantly higher in patients who lived in Chengdu, in patients who had longer dialysis vintage and in patients who went to the hospital more frequently (more than 3 times per week), as well as in patients who were more influenced by COVID-19 in terms of the severity of illness or daily life. We also found patients who had less confidence in overcoming the disease got higher IES scores (Table 5). Additionally, we did not find difference between IES and other variables, such as age, gender, education, dialysis modality, etc. (Table 5).

**Table 5.** IES scores of the study respondents.

|  | IES score [median (IQR*)] | P value |
| --- | --- | --- |
| **Gender** |  | 0.616 |
| Male | 8.00 (1.00-17.00) |  |
| Female | 8.00 (2.00-21.5) |  |
| **Age, years** |  | 0.097 |
| $\leq 40$ | 6.50 (1.00-14.00) | |
| 41-60 | 9.00 (3.00-23.00) |  |
| $\geq 61$ | 8.00 (0.00-14.00) | |
| **Marital status** |  | 0.268 |
| Married | 8.00 (2.00-19.00) |  |
| Single | 5.00 (1.00-13.00) |  |
| Divorced | 13.00 (5.00-26.00 ) |  |
| Widowed | 9.00 (0.00-24.75 ) |  |
| **Education** |  | 0.408 |
| Primary | 12.0 (1.50-28.75) |  |
| Junior | 8.00 (1.00-18.75) |  |
| Senior | 11.00 (3.00-18.25) |  |
| University degree or above | 7.00 (1.00-19.00) |  |
| **Occupation** |  | 0.762 |
| Medical staff | 3.00 (1.00-13.00) |  |
| Worker/farmer | 8.00 (1.50-15.50) |  |
| Teacher | 6.00 (1.00-11.00) |  |
| Government | 9.00 (3.75-27.25) |  |
| Company employee | 5.00 (1.00-19.25) |  |
| Retired | 11.00 (0.00-27.00) |  |
| Unemployment or others | 9.00 (2.00-20.00) |  |
| **Habitation** |  | 0.012 |
| City of Chengdu | 10.00 (3.00-23.00 ) |  |
| Other areas outside Chengdu | 6.00 (1.00-13.50 ) |  |
| **Dialysis vintage, years** |  | 0.006 |
| <1 | 5.00 (0.00-13.50 ) |  |
| 1-2 | 6.00 (1.00-16.25 |  |
| 3-5 | 8.00 (3.00-23.50 ) |  |
| >5 | 13.00 (5.00-23.00 |  |
| **Dialysis modality** |  | 0.02 |
| HD | 11.5 (3.00-25.00) |  |
| PD | 8.00 (1.00-15.00) |  |
| **Frequency to hospital per week** |  | 0.041 |
| 0 | 7.00 (1.00-14.00) |  |
| 1-2 | 10.50 (1.25-21.25) |  |
| ≥3 | 12.00 (3.00- 25.00) |  |
| **Influence on severity of illness** |  | 0.000 |
| No | 4.0 0(1.00-11.00 ) |  |
| Mild | 10.00 (2.00-19.00 ) |  |
| Moderate | 20.00 (8.75-26.25 ) |  |
| Severe | 21.50 (7.75-56.25) |  |
| **Influence on daily life** |  | 0.000 |
| No | 5.00 (0.00-11.00 ) |  |
| Mild | 7.00 (2.00-15.25 ) |  |
| Moderate | 14.00 (5.00-27.00 ) |  |
| Severe | 21.50 (11.75-40.5 ) |  |
| **Who went to the hospital** |  | 0.889 |
| Only myself | 8.5 (2.00-19.00) |  |
| Replaced by family members | 8.00 (2.00-19.00) |  |
| Accompanied by family | 8.00 (1.50-17.50) |  |
| **Confidence in overcoming the disease** |  | 0.000 |
| Full | 6.00 (1.00-15.00) |  |
| Moderate | 13.00 (7.250-23.00) |  |
| Rare | 27.00 (6.00-) |  |
| **Relationship with family** |  | 0.05 |
| Better | 10.00 (2.50-24.50) |  |
| Same | 7.00 (1.75-16.25) |  |
| Worse | 20.00 (7.5-28.50) |  |

\*IQR: Inter-quartile range

The significant factors above were analyzed for IES score using multivariate regression analysis. It was found that dialysis vintage, the impact of COVID-19 on the severity of illness or daily life, and confidence in overcoming the disease were independent risk factors (Table 6). Dialysis modality, frequency to hospital, habitation, whether acquired protective information from medical staff, relationship with family and other socio-demographic factors had no effect in the joint analysis.

**Table 6.** Results of multivariate analysis: risk of IES score.

| Model | Unstandardized |  | Standardized | Sig |
| --- | --- | --- | --- | --- |
|  | Coefficients |  | Coefficients |  |
|  | B | Std. Error | Beta |  |
| (constant) | 6.03 | 6.394 |  | 0.347 |
| Influence on state of illness | 3.68 | 1.370 | 0.195 | 0.008 |
| Dialysis vintage | 2.378 | 0.819 | 0.176 | 0.004 |
| Confidence in overcoming the disease | -4.309 | 1.849 | -0.145 | 0.021 |
| Influence on life | 2.84 | 1.361 | 2.087 | 0.038 |

## DISCUSSION

The outbreak of COVID-19 at the end of 2019, which spread rapidly and widely, brought great psychological pressure to the public [23]. In this cross-sectional study, we explored the psychological distress of patients undergoing dialysis and compared the difference between HD and PD during the lockdown period. We found that the median IES score for all the enrolled patients was 8.00 (2.00-19.00), which belonged to the subclinical dimension of post-traumatic stress symptoms. HD patients had significant higher IES scores than PD patients. And we observed that HD patients’ psychological reaction to stress was mainly avoidance. Our study also showed that dialysis vintage, the impact of COVID-19 on the severity of illness and daily life, and confidence in overcoming the disease were independent risk factors for IES.

We were surprised to find the median score of IES was not high in the whole enrolled patients. The median IES scores of HD and PD patients were 11.5 and 8.00, and represented as mild range and subclinical range of post-traumatic stress symptoms, respectively. Most of the dialysis patients could face the current epidemic calmly. Only 22.4% HD patients and 13.4% PD patients had the IES score more than 26, which indicated moderate or severe impact. We should pay more attention to these patients and provide sufficient psychological support in advance.

Several reasons might lead to higher IES scores and more severe trauma-related stress symptoms in HD patients. Firstly, PD patients could complete the dialysis treatment at home. And our study found that the frequency to hospital was much lower in PD patients. There were 80.1% of PD patients didn’t go to the hospital since the outbreak and 30.1% of them chose their family members to take the place of them for purchasing medicine or peritoneal dialysis fluid. On the contrary, every HD patient had to go to the dialysis center. Regular hemodialysis was very important [24] and was the main reason for going out in HD patients (93.4%). Additionally, in order to decrease the spread of COVID-19, the traffic control measures and community restrictions were adopted by the government [25]. Normal people were restricted from going out and medical proofs of dialysis treatment were required in some communities. This might cause some inconvenience for HD patients to go to dialysis centers. As we found out, 94.7% HD patients went to the hospitals at least 3 times a week. Significant higher frequency of going out for dialysis treatment and inconvenient transportation might put great psychological pressures on HD patients.

Besides, HD patients had more concerns of being infected and lack of protective equipment and might become more stressful. Hospitals were high-risk areas for infection because of the influx of febrile patients [26]. We found that dialysis treatment and buying necessaries were the main reasons of going out for HD patients and PD patients, respectively. People were advised to avoid going to such places during the epidemic [27]. Shopping can be avoided in a great extent, but dialysis is essential. Therefore, 44.7% HD patients worried about themselves being infected. In contrast, only 34.6% PD patients were anxious about this. Meanwhile, the shortage of masks and other protective materials was serious in February [28]. Higher frequency of going out required more protective materials. Of the patients we included, 48.7% HD patients concerned about the shortage of protective equipment, while only 28.2% PD patients worried about this. The lack of protective materials made HD patients bear greater psychological pressure. Based on all these factors, HD patients might have higher IES scores than PD patients. This probably also lead to more HD patients think that COVID-19 had an influence on their diseases or the severity of their diseases.

In the univariate analysis, we found that the frequency to hospitals was related to IES as well. The more often patients went to the hospitals, the higher risk of infection and more worries, which we had discussed above. Univariate analysis also showed HD might relate to higher IES scores. It was reported that HD patients might choose HD because of their poor independence [29]. For this reason, they might need more external support, which would become more obvious when they were confronted with such major public health events.

As for the habitation, univariate analysis found that people living in the city of Chengdu had higher IES scores. Chengdu is the capital of Sichuan Province and is also where our hospital located. A study investigated that PD uptake increased with increasing remoteness [30] and most of the hemodialysis patients lived in the urban areas [31]. The urban areas might have greater concentration and connectedness of people, which increased the possibility of infections [32]. In comparison, the patients living in other areas outside Chengdu were less worried about getting infected, probably because of the relatively small population.

After conducting the multivariate analysis for IES, our study suggested that dialysis vintage was the independent factor of IES. Precedent studies reported that dialysis patients with longer dialysis duration usually accompanied more comorbidities [33,34]. It was reported that higher illness severity might contribute to HD patients’ stress [29,35]. The longer dialysis vintage, the more severe of their illness, which might cause the patients become more distressed.

What’s more, we found that the influence on severity of illness and the influence on daily life were also the independent factors of IES. Worsening of the illness [29,35] and inconvenience of daily life might throw more stress to the dialysis patients.

Multivariate analysis also revealed that the confidence to overcome the disease was related to IES. Patients with less confidence got higher IES scores. Self-efficacy is the extent or strength of one’s belief in one’s own ability to complete tasks and reach goals [36]. And self-efficacy is one of the strongest predictors of anxiety in ESRD [37]. The patients who lacked confidence to overcome the disease might have lower self-efficacy and thought that they did not have the ability to fight against the virus. As a result, they might be more distress.

An interesting finding was the demands of psychological support were different according to the IES score and according to the patients’ requirements. Only 22.4% HD patients and 13.4% PD patients were classified as having moderate or sever post-traumatic stress symptoms, which need psychological support [22]. But there were 50% HD patients and 45.5% PD patients considered psychological support was necessary. More patients felt that they should get psychological support subjectively. This phenomenon might reflect these patients were trying to use the available resources to go through the epidemic.

Another interesting finding was although HD patients had higher IES scores, the demand for psychological support had no significant difference between the two groups. We think HD already received more psychological support and protective information from medical workers. Therefore, the demand for more psychological support had no difference in the two groups. But HD patients should still be given more psychological support. This could also be proven as HD patients were more inclined to relieve stress by chatting with medical staff than PD patients.

According to our study, we could see that psychological stress was inevitable among some dialysis patients. It is important to identify high-risk individuals and provide psychological intervention for them in advance. Previous studies about SARS pointed out that the psychological implications of the epidemic should not be ignored [38,39]. In order to relieve their stress, medical staff, including the physicians and nurses of dialysis center and professional psychologists as well, should offer psychological support as soon as possible.

Our study found about 60% PD patients hoped more convenient delivery service of drug. The hospitals might improve the delivery service during this epidemic. Some patients expected more flexible and convenient way for adjusting their treatment regimens. Remote medical treatment through the online service might be a good choice, particularly telephone-based and internet-based counseling [40].

There were several limitations in our study. Firstly, the sample size was relatively small. We only included 232 dialysis patients in total, especially the small number in HD patients. Furthermore, smart phones were used to conduct the questionnaire survey. The information of some elderly patients was not available because they could not use smart phones. This might lead to non-respondent bias in our study. Maybe it would be better to use telephone survey for the elderly patients in future study.

Until now, no precedent studies have reported on COVID-19-related stress for the dialysis patients. As far as we know, this is the first research comparing the psychological distress between HD and PD patients during the public health emergency. COVID-19 is still spreading worldwide and probably will last for a long period. Our study may have some practical significance for dialysis patients during this epidemic.

## CONCLUSION

This study explored the psychological distress of dialysis patients during the lockdown period of the epidemic of COVID-19. HD patients had significant higher IES scores and more severe trauma-related stress symptoms than PD patients. Dialysis vintage, confidence to overcome COVID-19, influence on state of illness and the influence on daily life were independent risk factors for IES. When major public health events occurred, careful psychological estimate and sufficient psychological support should be provided to the dialysis patients, especially to the HD patients.

## Data Availability

The data that support the findings of this study are available from the corresponding author upon reasonable request.

## Acknowledgment

There are no acknowledgments to declare.

## Statement of Ethics

This cross-section study was approved by the biomedical ethics committee of West China Hospital of Sichuan University (IRB NO.2020-185). And the informed consent was obtained for all patients.

## Disclosure Statement

The authors have no conflicts of interest to declare.

## Funding Sources

There are no funding sources to declare.

## Author Contributions

XXX, WXF, ZXL and LZ contributed to the design of the questionnaire. XXX, WXF, ZXL and PL contributed to the data collection. XXX, WXF, ZXL and ZZY contributed to the data analysis, interpretation, and manuscript preparation of this study. LZ contributed to the concept, design, data analysis, interpretation, and manuscript preparation and supervised of this study. All authors have read approved the final manuscript.

## Notes

### Competing Interest Statement

The authors have declared no competing interest.

